# Vaccine hesitancy strongly correlates with COVID-19 deaths underreporting

**DOI:** 10.1101/2022.02.27.22271579

**Authors:** Adam Sobieszek, Miriam Lipniacka, Tomasz Lipniacki

## Abstract

Vaccine acceptance is a key factor in achieving high immunization coverage and reducing the death toll of COVID-19 pandemic. Analyzing data from Europe and Americas we demonstrated that vaccine hesitancy strongly correlates with underreporting of COVID-19 deaths and cases. This correlation cannot be explained by the differences in economic indexes: GDP and Gini coefficient (measure of income inequalities). There is no correlation of vaccination percentage and Gini coefficient and the correlation with GDP is decreasing in time. The most striking is the comparison of Eastern European and South American countries; the latter group of countries shows significantly higher vaccination percentage while having a lower or comparable GDP and higher Gini coefficient. The analysis suggests that timely and reliable information about the COVID-19 cases and the associated deaths plays a key role in achieving population-wide vaccine acceptance.

## Introduction

Vaccinations were proved to be very effective in reducing the death toll and medical costs of COVID-19 pandemic^1–3^. Despite the fact that effectiveness of vaccines in reducing COVID-19 deaths was proved both by clinical trials and further studies during subsequent pandemic waves associated with new variants of concern^3^, vaccine hesitancy is still a widespread attitude in some countries. The vaccine acceptance and hesitancy were investigated in numerous survey studies in which people were asked about their willingness to vaccinate^4–7^.

In the early study (June, 2020), Lazarus et al. surveyed people in 19 countries, that comprises about 55% of world population, to find differences in willingness to vaccinate rates that range from more than 85% (in China and Brazil), about 75% in India and the US, to less than 60% (in France, Poland and Russia)^8^. The high willingness to vaccinate in China (85%), Brasil (88%) and UK (89%) and the low willingness to vaccinate in Russia (42%) was confirmed by a later (February 2021) IPSOS study^4^. Lindholt et al. studying Denmark, France, Germany, Hungary, Sweden, Italy, UK and USA between September 2020 to February 2021 found large variations in vaccine acceptance ranging from 83% in Denmark to 47% in France and Hungary^6^. However, according to the data retrieved from Our World in Data^5^ in France the significant reduction in vaccine hesitancy from 47% to 16% was observed during 2021, resulting in the actual 80% first dose vaccination rate (and nearly 50% booster rate) in the beginning on February 1, 2022.

Arce et al. reports a survey performed in the second half of 2020 across seven low-income countries (Burkina Faso, Mozambique, Rwanda, Sierra Leone and Uganda), five lower-middle-income countries (India, Nepal, Nigeria and Pakistan) and one in an upper-middle-income country (Colombia)^7^. The average acceptance rate across these countries was found equal 80.3% (95% CI 74.9–85.6%) considerably higher than in US 64.6% (95% CI 61.8–67.3%, high income) and Russia 30.4%, (95% CI 29.1–31.7%, upper-middle income).

In summary, the survey studies indicate high differences in vaccine acceptance, but without clear relation between the acceptance rate and the income *per capita*. People in China, Brazil and West European countries exhibited high willingness to vaccinate, while people in Eastern European countries (Russia, Poland, Hungary) had the lowest willingness. The survey studies are a natural method to investigate vaccine acceptance in the time period in which people do not have a full access to vaccines. When vaccines become widely available in a given country, however, the vaccination rate is a more straightforward measure of vaccination willingness, as it reports actual people behavior. As the vaccine availability correlates with the gross domestic product (GDP) *per capita* the vaccination rate reflects both the people’s willingness to vaccinate and the country’s economic standing. In European and American countries, considered in this study, the importance of the second factor is decreasing in time. Keeping in mind the possible influence of economic factors we will consider the proportion of people that obtained at least one vaccination dose as a measure of vaccine acceptance.

Surveys indicate that respondents reporting higher levels of trust in information from government sources were more likely to accept a vaccine^6,8^. It is thus important to analyze how reliability of COVID-19 pandemic reporting correlates with vaccination rates and so vaccine acceptance. Numerous studies indicate that in significant proportion countries the number of excess deaths is substantially higher than the number of COVID-19 confirmed deaths, suggesting a significant COVID-19 deaths underreporting^9,10^. Similarly, the ratio of cumulative COVID-19 confirmed cases to cumulative COVID-19 associated deaths (defined as the maximum of the COVID-19 confirmed deaths and excess deaths) varies significantly between countries (also before the vaccination campaign) suggesting underreporting of COVID-19 cases. This indicates that some countries report only a relatively small fraction of COVID-19 deaths and cases^9^.

We use the COVID-19 deaths and cases reporting as a measure of reliability of disclosed information about epidemic and show that it strongly correlates with vaccination rates in Europe and Americas. Then focusing on Eastern Europe and South America we show that generally higher vaccination rates and more reliable epidemic reporting in the second region may not be explained by GDP PPP (purchase power parity) *per capita* or Gini coefficient (that measures economic inequalities with a country).

## Results

We analyze the relation between the reliability of COVID-19 deaths reporting and the vaccination rates in Europe and Americas. As measure of reliability of the COVID-19 death reporting we chose the ratio *F* of the cumulative COVID-19 confirmed (and reported) deaths to the cumulative excess deaths, both calculated since the country’s first 50 COVID-19 confirmed deaths until a given time *t*_deaths_, based on the Economist database^10^. Intuitively, the highest is the ratio *F*, the better is COVID-19 death reporting. In most analyzed countries *F <* 1, there are however some countries for which *F >* 1, as well as three countries (Bahamas, Norway, Uruguay) with negative numbers of excess deaths. It is important to note that Australia (between April 6, 2020 and November 28, 2021) and New Zealand (between February 3, 2020 and January 30, 2022), not considered in this study, despite having small numbers of COVID-19 confirmed deaths, have overall negative excess of mortality equal to 0,05%^10^. This is considered to be a consequence of non-medical anti-epidemic interventions that reduced deaths associated with other infectious diseases such as influenza^11^. The possible reduction of non-COVID-19 deaths suggests that in countries with reliable reporting, in which nearly all deaths associated with COVID-19 infections are reported, *F* may exceed 1. However, the increase of *F* to the values much greater than 1 is unlikely associated with further reporting improvement.

In Fig. 1a, we thus assume an arbitrary threshold (varied in Fig. 1e) *F*_threshold_ = 1.2 and assign it to countries for which *F* > *F*_threshold_ or to countries for which the number of excess deaths is negative. In order to measure vaccine acceptance rather than the availability of vaccines in considered countries we defined the vaccination rate *S* as the share of individuals that received at least one vaccination dose before a given time *t*_vacc_. For Fig. 1a we set *t*_deaths_ = March 31, 2021 and *t*_vacc_ = September 30, 2021, to find that *ρ*_S-F_, the population-weighted Pearson correlation between *S* and *F*, is *ρ*_S-F_ = 0.78. To support this finding we calculate also *ρ*^*^_S-F_, the population-weighted Pearson correlation between rank values of *S* and *F*, and obtain *ρ*^*^_S-F_ = 0.79. Correlation between rank values is an appropriate measure of relation also between variables that not are not normally distributed. Of note, the non-weighted Pearson correlation on ranks is known as Spearman correlation. Our choice of *t*_deaths_ < *t*_vacc_ allow to study the effect of deaths reporting on vaccine acceptance, simultaneously minimizing the effect of vaccinations on excess deaths. The analyzed countries are shown on the map in Fig. 1b in which colors indicate geographical regions, while gray areas indicate countries with insufficient reporting to be included in Fig. 1a (see Methods for details).

**Figure 1.**
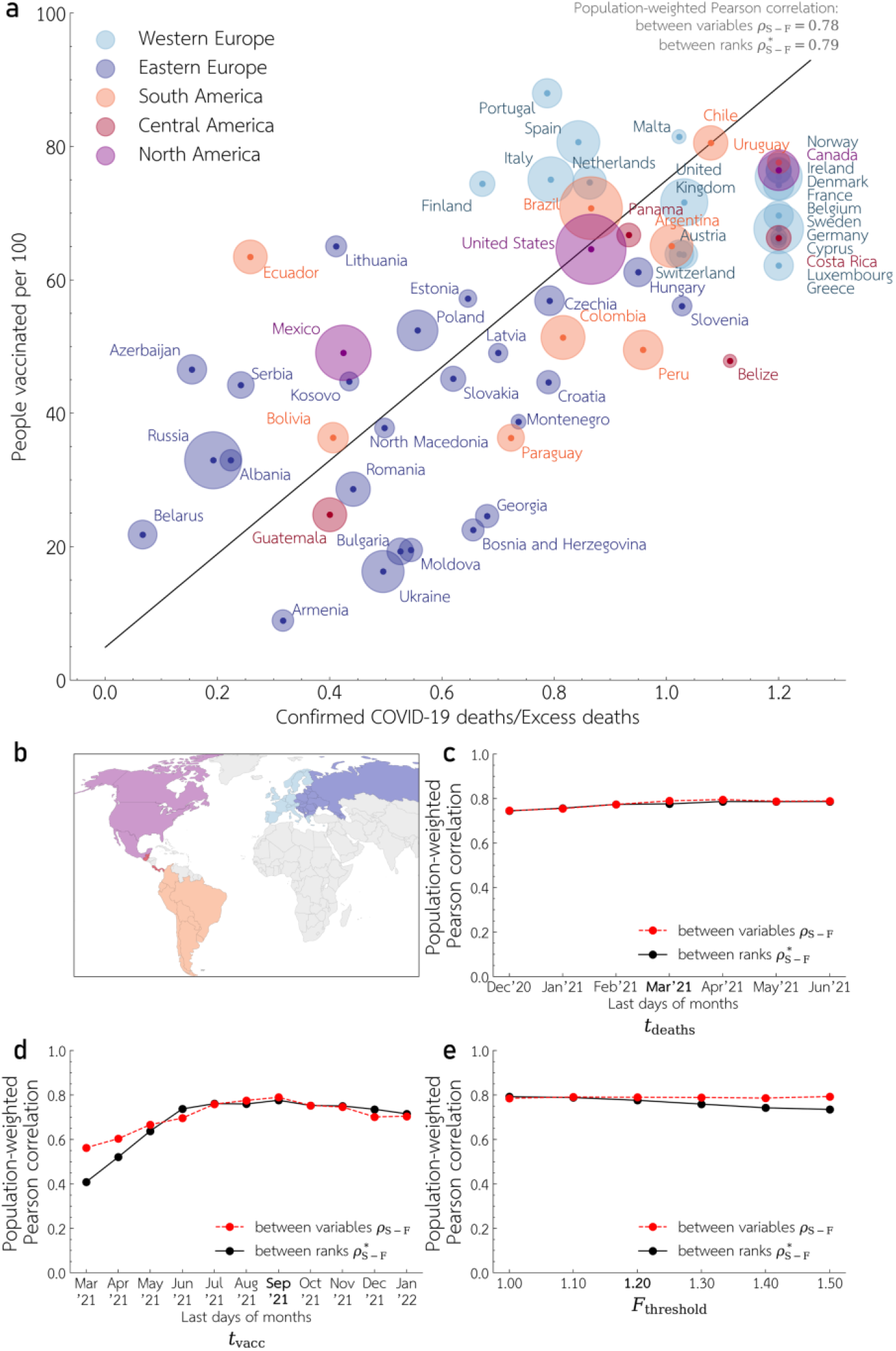
Relation between COVID-19 deaths reporting and vaccination rate in Europe and Americas. (**a**) Vaccination rate (share of people that received at least one dose), *S*, till *t*_vacc_ = September 30, 2021 versus the ratio of cumulative COVID-19 confirmed deaths to cumulative excess deaths, *F*, calculated since country’s first 50 COVID-19 confirmed deaths until *t*_deaths_ = March 31, 2021. For countries with negative excess deaths, or in which *F* > *F*_threshold_ =1.2, *F* is assumed equal *F*_threshold_. Disk area of each country is proportional to the square root of its population; the linear trend is indicated by black line. (**b**) Map of the five considered geographical regions indicated by colors; gray areas denote countries with insufficient reporting to be included in the analysis. (**c**), (**d**) and (**e**) Population-weighted Pearson correlation coefficients *ρ*_S-P_ between *S* and *F*, and *ρ*^*^_S-P_ between rank values of *S* and *F* as a function of *t*_deaths_ in (**c**), *t*_vacc_ in (**d**) and *F*_threshold_ in (**e**). Nominal values of *t*_deaths_, *t*_vacc_ and *F*_threshold_ used for Panel (**a**) are in bold on *x*-axises in Panels (**c**), (**d**), and (**e**).

Since the nominal values of *t*_deaths_, *t*_vacc_ as well as *F*_threshold_ chosen for Fig. 1a, are to some extent arbitrary in Figs. 1c, 1d and 1e we analyze dependence of *ρ*_S-F_ as well as *ρ*^*^_S-F_ on these three parameters. Generally, we may notice that *ρ*_S-F_ is close to *ρ*^*^_S-F_ indicating linear relation between *S* and *F*. Both *ρ*_S-F_ and *ρ*^*^_S-F_ weakly depend on *t*_deaths_ (in range December 31, 2020 - July 30, 2021) and on *F*_threshold_ (in range 1.0-1.5). Not surprisingly both correlation coefficients grow with *t*_vacc_ between April 30, 2021 and August 31, 2021 (i.e. during the most intense vaccination campaign), and stabilize between August 31, 2021 and January 31, 2022.

Reporting of COVID-19 cases (in addition to reporting deaths associated with COVID-19 infections) is the second natural measure of reliability of the epidemic reporting. In the early epidemic phase (till March 31, 2021), when the life-saving vaccines were not widely available, the ratio *P* of the cumulative COVID-19 confirmed cases to COVID-19 associated deaths is a good measure of reporting reliability. As shown in the Supplementary Figure 1, this ratio ranges from 5-10 for Mexico, Peru or Russia to about 100 for Denmark, Cyprus, Norway and Uruguay, suggesting that in the first group of countries only a tiny fraction of cases was confirmed and reported. The population-weighted correlation between *S* and *P* for *t*_cases_ = *t*_deaths_ = March 31, 2021 and *t*_vacc_ = September 30, 2021 is *ρ*_S-P_ = 0.63, and between rank values of *S* and *P* is *ρ*^*^_S-P_ = 0.64 (Supplementary Figure 1a). Both *ρ*_S-P_ and *ρ*^*^_S-P_ are roughly stable with respect to *t*_cases_ = *t*_deaths_ in range December 31, 2020 - June 30, 2021 (Supplementary Figure 1b) as well as with respect to *t*_vacc_ in the range March, 2021 - January 31, 2022 (Supplementary Figure 1c).

The correlations reported in Fig. 1 and Supplementary Figure 1 suggest that vaccine acceptance increases with reliability of epidemic reporting. It is natural to expect this correlation to result from some confounding factors such as differences of GDP *per capita* or Gini coefficient. Intuitively, the highly developed countries may afford both for the reliable epidemic reporting and swift vaccinations. We observe, however, that for *t*_vacc_ = September 30, 2021 the population-weighted Pearson correlation between GDP PPP (purchase power parity) *per capita* and vaccination rate is *ρ*_S-GDP_ = 0.41 (and between corresponding values, *ρ*^*^_S-GDP_ = 0.40), Fig. 2a, i.e. substantially lower than *ρ*_S-F_ (Fig. 1a). As expected, correlation *ρ*_S-GDP_ was very high in the early stage of the vaccination campaign (exceeding 0.8 between March 31 and June 30, 2021), when only the most wealthy countries of Europe and Americas had access to vaccines. The correlation dropped, however, below 0.33 after October 31, 2021 (Fig. 2c) when the vaccination rate started to reflect the population’s willingness to vaccinate, rather than access to vaccines. The additional observation is that all South America’s countries are above the trend line (indicating a relatively high vaccination rate with respect to GDP PPP *per capita*), while the majority of Eastern European countries are below the trend line (indicating relatively low vaccination rate with respect to GDP PPP *per capita*). Juxtaposing vaccination rate and Gini coefficient (Fig. 2b and Fig. 2d) suggests no correlation between them, but allows us to distinguish three groups of countries: Eastern European with low Gini coefficients and low vaccination rates, West European (and Canada) with similarly low Gini coefficients and significantly higher vaccination rates and American (excluding Canada) with a high Gini coefficient and diverse vaccination rates.

**Figure 2.**
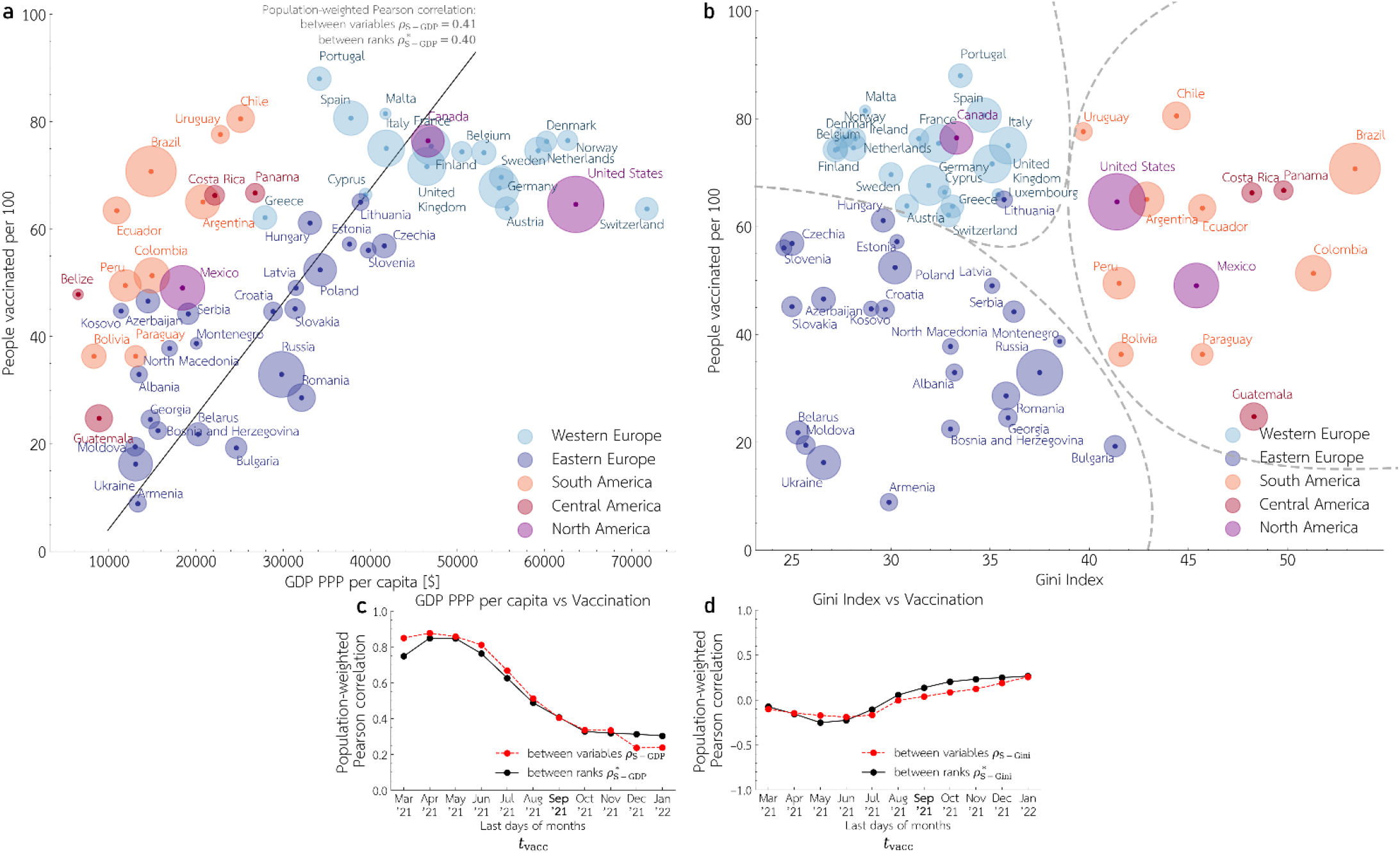
Relation between economic indexes and vaccination rate in Europe and Americas. (**a**) Vaccination rate, *S*, till *t*_vacc_ = September 30, 2021 versus GDP PPP *per capita*. Disk area of each country is proportional to the square root of its population; the linear trend is indicated by black line. (**b**) Vaccination rate, *S*, till *t*_vacc_ = September 30, 2021 versus Gini coefficient. (**c**) and (**d**) Population-weighted Pearson correlation coefficients *ρ*_S-GDP_ (and *ρ*^*^ _GDP_) between *S* and GDP PPP *per capita* (and the corresponding rank values) in (**c**) and *ρ*_S-Gini_ (and *ρ*^*^_S-Gini_) between *S* and Gini coefficient (and the corresponding rank values) in (**d**) both as a function of *t*_vacc_. Nominal value of *t*_vacc_ used for Panel (**a**) is in bold on *x*-axises in Panels (**c**), and (**d**).

The analysis in Fig. 2 suggests that higher vaccination rate may be explained by the higher GDP PPP only in the early phase of vaccination campaign when in part of the considered countries the vaccines were not widely available. Surprisingly, the vaccination rate is not anti-correlated with Gini coefficient; i.e. in the considered countries the economic inequalities are not associated with the lower vaccination rates. In further analysis we focus on South American and Eastern European countries that show surprising differences in vaccination rates.

In Fig. 3 we repeat the analysis from Fig. 1 restricting to these two geographical regions, obtaining for nominal *t*_deaths_ = March 31, 2021 and *t*_vacc_ = September 30, 2021, similar value of population-weighted Pearson correlation *ρ*_S-F_ = 0.75 (and *ρ*^*^_S-F_ = 0.8), Fig. 3a. The values of *ρ*_S-F_ and *ρ*^*^_S- F_ weakly depend on *t*_deaths_ (Fig. 3b), but significantly increases with *t*_vacc_ in consequence of the increasing availability of vaccines in South America in the considered period. Also, restricting to South American and Eastern European countries we observe a similar value of the weighted Pearson correlation between reporting of COVID-19 cases and vaccination rate, *ρ*_S-P_ = 0.62 (and *ρ*^*^_S-P_ = 0.67), see Supplementary Figure 2. Fig. 3 and Supplementary Figure 2 indicate that a higher vaccination rate in South America with respect to Eastern Europe correlates with the better reporting of COVID-19 deaths as well as of COVID-19 cases.

**Figure 3.**
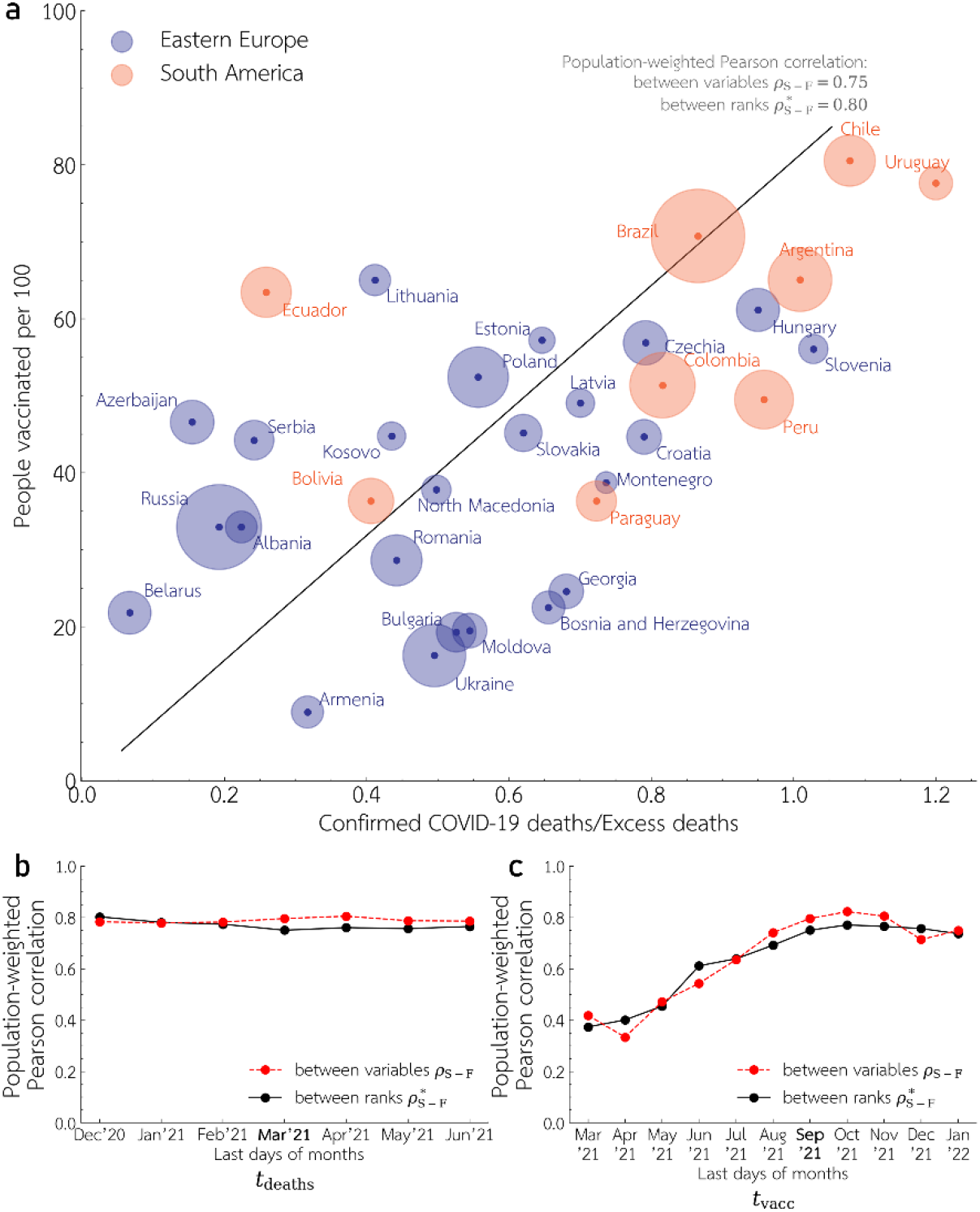
Relation between COVID-19 deaths reporting and vaccination rate in Eastern Europe and South America. **(a**) Vaccination rate, *S, t*v_acc_ = September 30, 2021 versus the ratio of cumulative COVID-19 confirmed deaths to cumulative excess deaths, *F*, calculated from the country’s first 50 COVID-19 confirmed deaths until *t*_deaths_ = March 31, 2021. Disk area of each country is proportional to the square root of its population; the linear trend is indicated by black line. (**b**) and (**c**) Population-weighted Pearson correlation coefficients *ρ*_S-F_ (and *ρ*^*^_S-F_) between *S* and *F* (and between rank values of *S* and *F*) as a function of *t*_deaths_ in (**b**), and *t*_vacc_ in (**c**). Nominal values of *t*_deaths_ and *t*_vacc_ used for Panel (**a**) are in bold on *x*-axises in Panels (**c**), (**d**).

Comparison of South America and Eastern Europe is important because a higher vaccination rate in South America with respect to Eastern Europe may not be explained by the higher GDP PPP *per capita*. As shown in Fig. 4a, although in each region, separately, vaccination rate increases with GDP, when both regions are considered jointly the population weighted Pearson correlation *ρ*_S-GDP_ and *ρ*^*^_S-GDP_ becomes negative for *t*_vacc_ ≥August 31, 2021 (Fig. 4c). The weighted correlation is strongly influenced by the two most populous countries: Russia and Brazil. As of September 30, 2021, Russia having GDP PPP *per capita* more than twice higher than Brazil had more than twice lower vaccination rate (Fig. 4a). Strikingly, in the epidemic period till September 30, 2021, 87% of excess deaths in Brazil were reported as COVID-19 deaths, while this proportion is 19% in Russia (Fig. 3a). Similarly, the ratio *P*(*t*_cases_ = *t*_deaths_ = September 30, 2021) of COVID-19 confirmed cases to COVID-19 associated deaths exceeded 30 in Brazil and was less than 10 in Russia (Supplementary Figure 2a), showing a much more reliable epidemic reporting in Brazil than in Russia. Finally, we may notice that all South American countries which on average have higher vaccination rates than Eastern European countries have also higher Gini coefficients, which results in surprisingly positive correlation between these two variables (Fig. 4b and Fig. 4d). Higher Gini coefficient and lower GDP PPP *per capita* in South America in relation to Eastern Europe rules out the possibility that higher vaccination rate and more reliable epidemic reporting are jointly a consequence of better economic standing of South America with respect to Eastern Europe.

**Figure 4.**
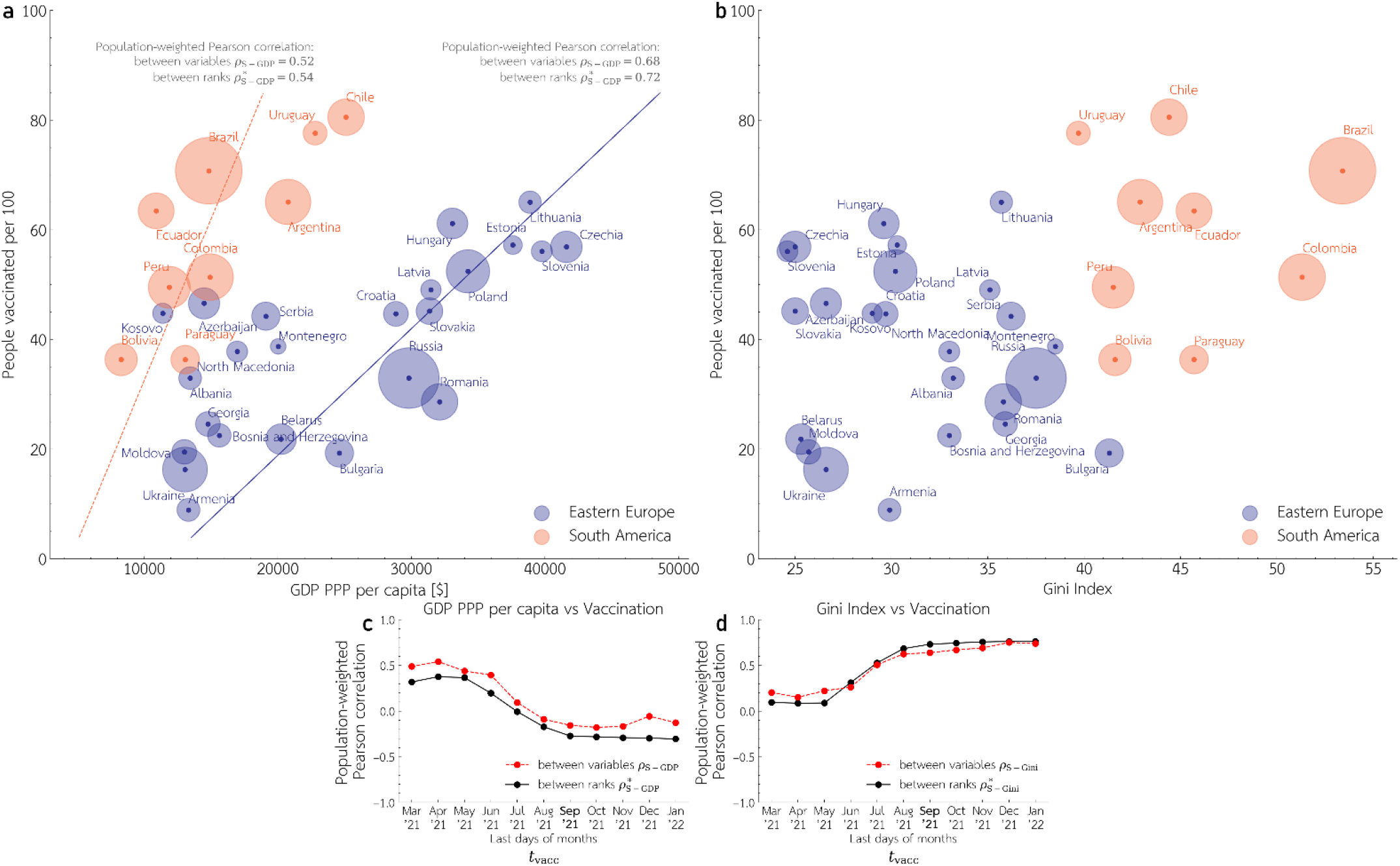
Relation between economic indexes and vaccination rate in Eastern Europe and South America. Vaccination rate, *S*, till *t*_vacc_ = September 30, 2021 versus GDP PPP. Disk area of each country is proportional to the square root of its population; the linear trends for Eastern Europe and South America are indicated by solid blue and dashed orange lines, respectively. (**b**) Vaccination rate, *S*, till *t*_vacc_ = September 30, 2021 versus Gini coefficient. (**c**) and (**d**) Population-weighted Pearson correlation coefficients *ρ*_S-GDP_ (and *ρ*^*^_S-GDP_) between *S* and GDP PPP *per capita* (and the rank values) in (**C**) and *ρ*_S-Gini_ (and *ρ*^*^_S-Gini_) between *S* and Gini coefficient in (**d**) both as a function of *t*_vacc_. Nominal value of *t*_vacc_ used for Panel (**a**) is in bold on *x*-axises in Panels (**c**), and (**d**).

## Discussion

Considering European and American countries we showed a significant correlation between reliability of COVID-19 pandemic reporting and vaccine acceptance. The reliability of reporting is assessed based on two measures: ratio, *F*, of cumulative COVID-19 confirmed cases to excess deaths (in the same period of time) and ratio, *P*, of COVID-19 confirmed cases to COVID-19 associated deaths. In some countries *F* is close to 1, and *P* is of order 100 (which suggests reliable reporting), while in some countries *F* is below 0.25, and *P* is below 10, which suggests that only a small fraction of COVID-19 deaths and cases are confirmed and disclosed to the public. Our measure of vaccine acceptance is the proportion of the population that received at least one vaccination dose. Obviously, the vaccinated proportion reflects both willingness to vaccinate, and availability of vaccines; however, as the vaccines become more and more available the first factor becomes dominant.

Intuitively, correlation between reliability of reporting and vaccinated proportion can be a consequence of confounding factors related to economic standing. High GDP and low Gini coefficient can positively influence both epidemic reporting and vaccination rate as the richer countries have more resources for COVID-19 testing and their citizens have better access to vaccines. In fact, there exists correlation between GDP PPP *per capita*, however this correlation (for European and American countries) decreases below 0.33 after October 31, 2021, when vaccines become widely available in Americas and Europe. Surprisingly, we observe no negative correlation between vaccination rate and Gini coefficient. This implies that high correlation between reliability of reporting and vaccinated proportion may not be fully explained by the economic factors.

The most striking is the comparison of South American and Eastern European countries. For this selected subset of countries, we observe a high population-weighted Pearson correlation between vaccination rate and COVID-19 deaths reporting, *ρ*_S-F_ = 0.75 (and population-weighted Pearson correlation between rank values of *S* and *F, ρ*^*^_S-F_ = 0.80) that may not be explained by economic factors. On average South American countries have a higher vaccination rate, lower GDP PPP *per capita*, and substantially higher Gini coefficients than Eastern European countries. Better economic standing of Eastern European countries, suggest that vaccine hesitancy in these countries follows from unreliable and insufficient reporting of COVID-19 pandemic. In the case of low mortality diseases such as COVID-19, people may not base their risk assessment on information from their relatives or friends, instead they require reliable information about epidemic risk from medical or governmental authorities. Comparison of South American and Eastern European countries suggests that underreporting of COVID-19 deaths and cases contributes to vaccine hesitancy. The vaccine hesitancy in turn has severe medical consequences; as of February 1, 2022 (see Methods for details), 12 out 15 countries with the highest cumulative excess deaths percentage, ranging from about 0.4% to 0.9%, are in Eastern Europe^10^.

## Methods

### Data Sources

The cumulative numbers of COVID-19 confirmed deaths and excess deaths were retrieved from the Economist repository^10^. Vaccination rates (defined as the proportion of people that received at least one dose) and the daily numbers of COVID-19 confirmed cases were retrieved from Our World in Data repository^12^. GDP PPP *per capita* (as of 2020) and Gini coefficients (in years 2017-2020, for all countries except Bosnia and Herzegovina, Germany, Guatemala and Montenegro for which last time of reporting was between 2011-2016) were retrieved from the World Bank repository^13,14^.

### Estimation of ratio of cumulative COVID-19 confirmed deaths to cumulative excess deaths *F*

The ratio *F* was estimated based on data from the Economist repository^10^ for last days of months from December 2020 to June 2021. For countries with weekly deaths reporting the cumulative numbers of COVID-19 confirmed deaths and excess deaths were retrieved from the last Sunday of the month (instead of the last day of the month). Countries for which cumulative excess deaths are not available for December 31, 2020 (the beginning of the period analyzed in Figs. 1c and 2b) were excluded from the analysis (see map in Fig. 1b). For four countries for which the latest reporting date is between December 31, 2020 and June, 30, 2021 the data from this latest reporting date is used also to estimate the ratio *F* in subsequent months. For Argentina, Belize and Costa Rica the latest data is December 31, 2020 while for Belarus the latest date is March 31, 2021.

### Estimation of the vaccination rate

In the case of Bulgaria and Moldova, we interpolated the missing data in some months based on adjacent data points. Albania started reporting vaccinations only in May of 2021 (long after the cumulative number of vaccinations exceeded 50), it is thus not included in correlations for *t*_vacc_ = March 2021 and *t*_vacc_ = April 2021. Bosnia and Herzegovina which started reporting vaccinations in April 2021, are not included in correlations for *t*_vacc_ = March 2021. Romania stopped reporting first dose vaccinations after September 27, 2021, it is thus not included in correlations for *t*_vacc_ = October 2021-January 2022.

### Estimation of excess deaths till February 1, 2022

For countries that do not report excess deaths till February 1, 2022, we estimate this number by assuming that the ratio *F* of cumulative COVID-19 confirmed deaths to excess deaths is constant in time. The accumulated excess deaths are calculated by dividing the accumulated confirmed deaths by the country specific ratio *F* from the last date available. This estimation is used only to conclude that 12 out of 15 countries with the highest excess deaths per million are from Eastern Europe.

## Data Availability

All data produced in the present work are contained in the manuscript

## Data availability

All data used in this study are referenced. COVID-19 mortality datasets (excess deaths and COVID-19 confirmed deaths) were retrieved from Economist repository (https://github.com/TheEconomist/covid-19-excess-deaths-tracker). Vaccination rates and daily numbers of COVID-19 cases were retrieved from Our World in Data repository (https://github.com/owid/covid-19-data), GDP PPP *per capita* and Gini coefficients were retrieved from World Bank repository (https://data.worldbank.org/indicator).

## Author contributions

Conceptualization, Adam Sobieszek, Miriam Lipniacka, Tomasz Lipniacki; Data curation and Software, Adam Sobieszek; Investigation, Adam Sobieszek and Miriam Lipniacka; Writing, Adam Sobieszek, Miriam Lipniacka, Tomasz Lipniacki;

## Acknowledgments

This study was supported by the National Science Centre Poland OPUS, grant 2018/29/B/NZ2/00668. The funding agency had no role in study design, data collection, and analysis, decision to publish, or preparation of the manuscript.

## Supplementary Figures

**Supplementary Figure 1.**
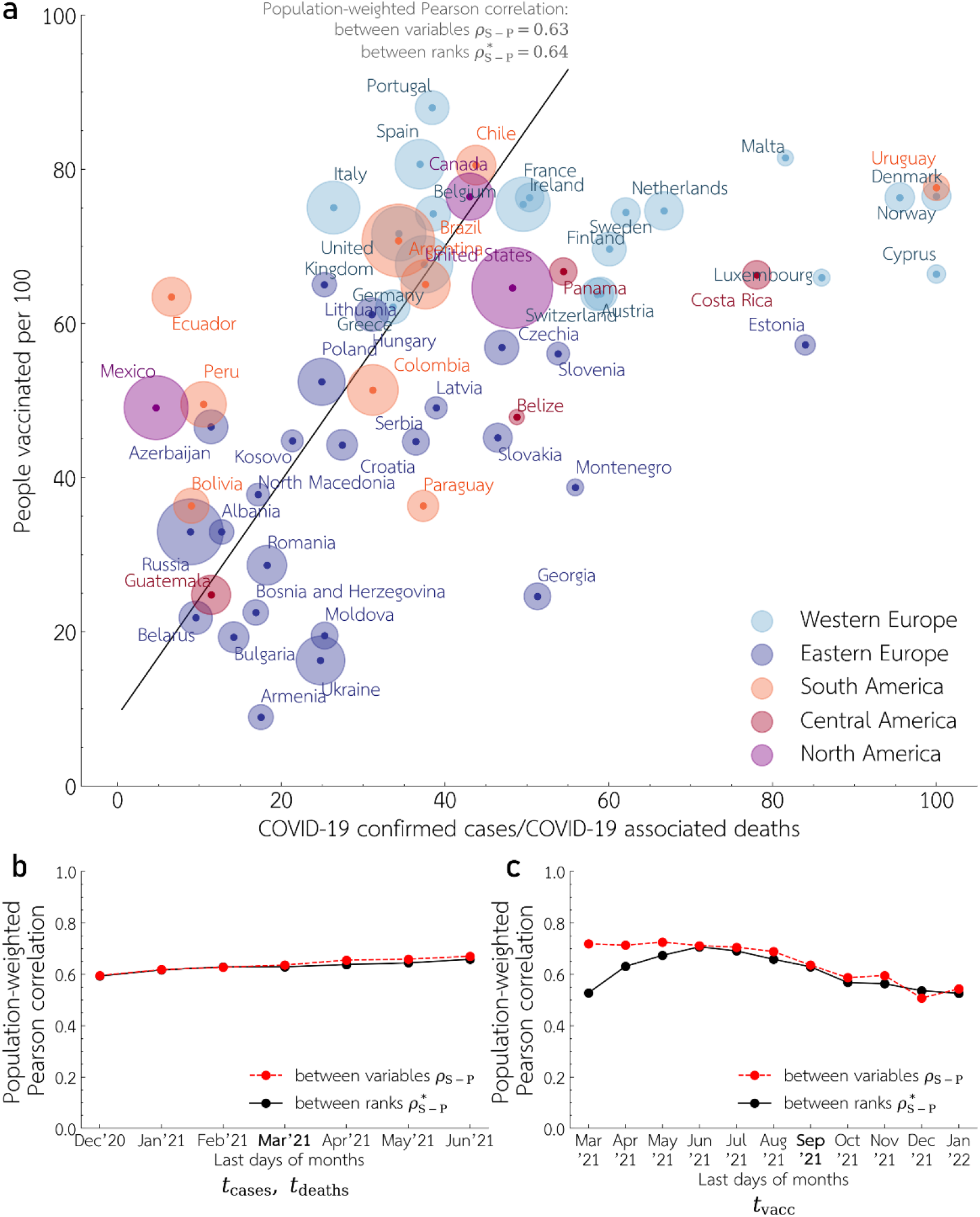
Relation between COVID-19 cases reporting and vaccination rate in Europe and Americas. (**a**) Vaccination rate, *S*, till *t*_vacc_ = September 30, 2021 versus the ratio *P* of cumulative COVID-19 confirmed cases to COVID-19 associated deaths (defined as the maximum of the cumulative COVID-19 confirmed deaths and cumulative excess deaths). The ratio *P* is calculated based on data accumulated since the country’s first 50 COVID-19 confirmed deaths until *t*_cases_ = *t*_deaths_ = March 31, 2021. Disk area of each country is proportional to the square root of its population; the linear trend is indicated by black line. (**b**) and (**c**) Population-weighted Pearson’s correlation coefficients between *ρ*_S-P_ (and *ρ*^*^_S-P_) between *S* and *P* (and between rank values of *S* and *P*) as a function of *t*_deaths_ in (**b**), and *t*_vacc_ in (**c**). Nominal values of *t*_cases_ = *t*_deaths_ and *t*_vacc_ used for Panel (**a**) are in bold on *x*-axises in Panels (**c**), (**d**).

**Supplementary Figure 2.**
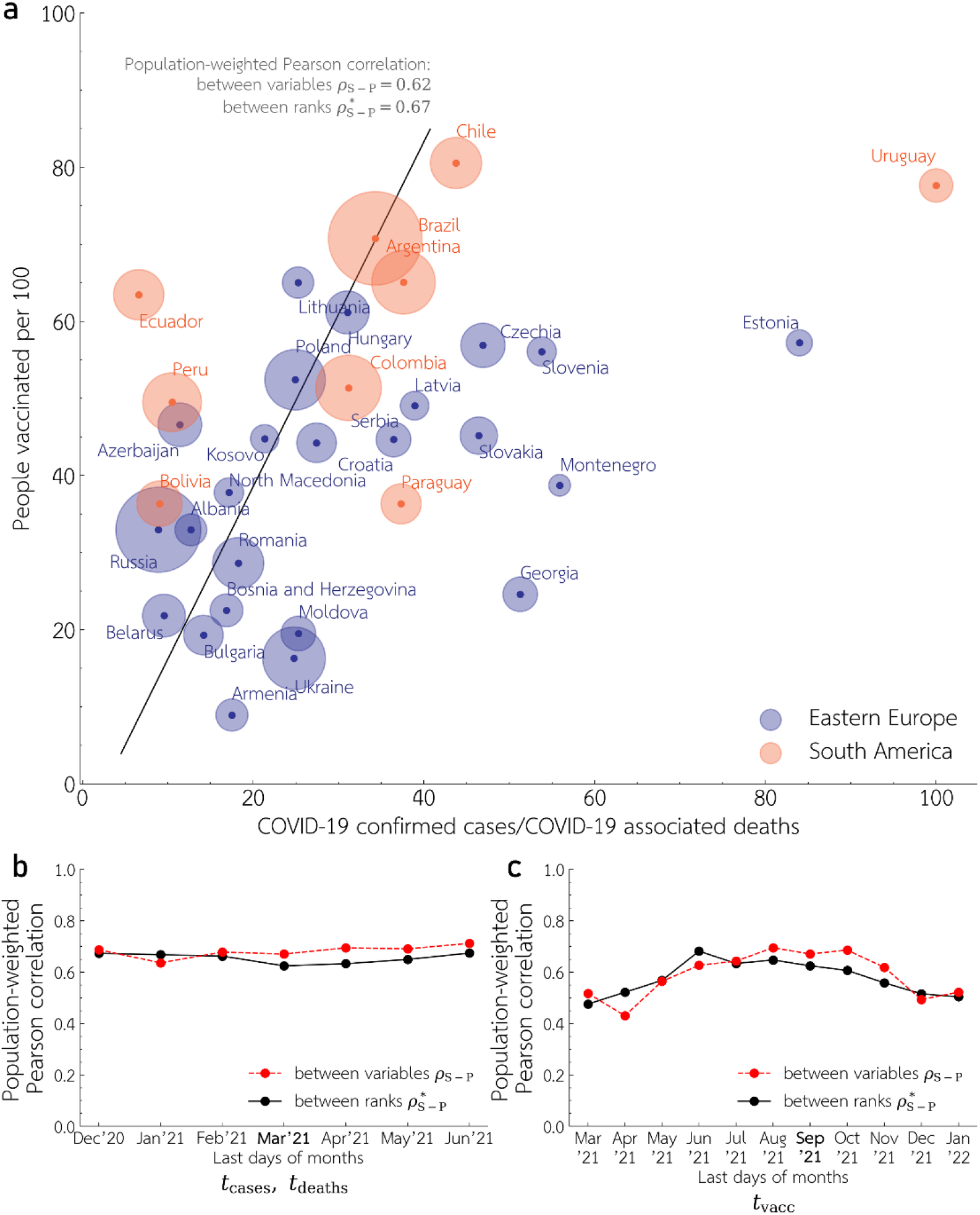
Relation between COVID-19 cases reporting and vaccination rate in Eastern Europe and South America. (**a**) Vaccination rate, *S*, till *t*_vacc_ = September 30, 2021 versus the ratio *P* of cumulative COVID-19 confirmed cases to COVID-19 associated deaths (defined as the maximum of the cumulative COVID-19 confirmed deaths and cumulative excess deaths). The ratio *P* is calculated based on data accumulated since the country’s first 50 COVID-19 confirmed deaths until *t*_cases_ = *t*_deaths_ = March 31, 2021. Disk area of each country is proportional to the square root of its population; the linear trend is indicated by black line. and (**c**) Population-weighted Pearson’s correlation coefficients between *ρ*_S-P_ (and *ρ*^*^_S-P_) between *S* and *P* (and between rank values of *S* and *P*) as a function of *t*_deaths_ in (**b**), and *t*_vacc_ in (**c**). Nominal values of *t*_cases_ = *t*_deaths_ and *t*_vacc_ used for Panel (**a**) are in bold on *x*-axises in Panels (**c**), (**d**).

